# A Study on the Understanding and Impact of Life Education on End-of-Life Concepts among the Elderly

**DOI:** 10.1101/2025.08.06.25332589

**Authors:** Tzu-Yu Chou

## Abstract

**Background/Objectives:** After entering a super-aged society, the clarity of self-worth and beliefs becomes particularly important for the elderly when facing illness and death. Life education can effectively assist in decision-making at the end of life, enhancing quality of life and dignity. In 2025, Taiwan will enter a super-aged society, and the government has introduced the Patient Autonomy Law to promote patient autonomy through Advance Care Planning (ACP) in specific clinical situations, aiming to achieve a dignified end of life. However, challenges remain in the promotion process. Previous research on medical autonomy and end-of-life concepts has primarily focused on terminal cancer patients and their families. This study aims to fill the gap in the literature regarding the perspectives of healthy older adults on a dignified end of life and to provide references and recommendations for future life education targeting this group.

**Methods:** This study employs a quasi-experimental design and utilizes a cross-sectional survey method. It analyzes older learners aged 55 and above in southern Taiwan (including Tainan City, Kaohsiung City, and Pingtung County), who are randomly assigned to experimental and control groups. The intervention involves multimedia teaching to assess their understanding and acceptance of end-of-life concepts. A shortened version of the Miyoshi questionnaire is used to evaluate these concepts, combined with items related to medical autonomy for older adults, with pre- and post-intervention measurements. Data processing involves repeated measures t-tests and analysis of covariance, with data analysis conducted using SPSS 22.0. The research instruments underwent item analysis and exploratory analysis.

**Results:** This study explores the impact of an intervention program on sen-ior learners’ perceptions of end-of-life concepts and medical autonomy. The results in-dicate that the experimental group experienced significant improvements in pre- and post-test scores across four dimensions: peaceful passing, informed consent, death awareness, and medical autonomy, with particularly notable effects in the areas of peaceful passing and death awareness. The research highlights that “disclosure of medical conditions” and “doctor-patient communication,” through a biomedical model or empirical principles, can effectively enhance doctor-patient interactions. Quantita-tive pre- and post-test scores demonstrate significant outcomes in clinical practice.

**Conclusion:** Facing the challenges of a super-aged society, the desire of the elderly for autonomy in choice and dignified death is increasingly growing. This study explores the concept of end-of-life among older learners in southern Taiwan. The results indicate that multimedia teaching provides a flexible learning model that enhances learners’ sense of participation. Through interaction and visual design, older adults can more effectively grasp and deeply understand complex concepts such as medical autonomy, dignified death, psychological preparedness, and quality of life. This not only addresses the challenges of promoting medical autonomy but also provides references for future policy formulation concerning the elderly.

## Introduction

The demands of older adults for health care services are more diverse. Especially in a super-aged society, there has been a significant increase in emergency medical care, palliative care, and end-of-life services (Li Fang-nian et al., 2018). Despite advancements in medical technology, clarifying self-worth and beliefs becomes even more important for older adults in facing the challenges of illness and death. Life education can effectively assist them in making good end-of-life decisions and enhance their dignity in dying (Hsu He-ling, 2020).In the past, Taiwan protected the end-of-life medical rights of terminally ill patients through the “Hospice Palliative Care Act.” However, whether this approach can enable patients to make end-of-life decisions based on self-awareness and informed consent remains a question (Chen Shipei et al., 2019). Therefore, effectively utilizing the right to refuse medical treatment and addressing the conflicts among patient autonomy, family values, and medical ethics in end-of-life care is crucial for avoiding ethical dilemmas caused by ineffective treatments (Cai Changying, 2020). In response to this, the Taiwanese government introduced the Patient Autonomy Act, which promotes dignified end-of-life care through Advance Care Planning (ACP) and Advance Directives (AD) in specific clinical situations, thereby applying medical autonomy. This legal framework aims to further improve the doctor-patient relationship (Lin Zhengpei, Peng Rengui, 2019).

However, in clinical practice, challenges in decision-making arise when patients must make choices regarding unknown medical conditions. Additionally, physicians often face difficulties in assessing prognosis and lack effective communication skills, which leads to a cognitive gap between doctors and patients in their understanding (Huang Junjie, 2019). Furthermore, in American jurisprudence, Advance Directives (AD) have not been clearly established as the primary means of realizing patient autonomy (Zhang Zhaotian, 2024). When confronted with end-of-life decisions, whether patients can make clear choices (Liao Yingjin et al., 2022) often differs from the decision-making patterns exhibited by their family members, leading to discrepancies and even contradictions between the two (Wu Jiaying et al., 2020). If we can deepen our understanding of holistic health through death literacy-related life education and clinical evidence, incorporating an integrated teaching model that includes the physical, mental, and spiritual aspects (Liu Yizhai, 2018), we can explore and impart the skills and communication abilities necessary for end-of-life care (Ma Ruiju et al., 2020). By doing so, we can further assist individuals in achieving a fulfilled sense of death and life meaning under the vision of “comfort for the living, peace for the departed” (Liu Yizhai, Liu Shimia, 2019).

In modern times, with the development of information technology, utilizing multimedia teaching models can provide convenient and flexible resources, creating interactive frameworks (Liao et al., 2019). This approach offers rich and in-depth knowledge while adhering to principles of dynamism, consistency, and excess, thereby providing diverse learning opportunities. It can effectively enhance learners’ concentration, reduce interaction pressure, and alleviate fatigue, promoting humane self-regulated learning and experiences for the elderly (Wei, 2019). This, in turn, strengthens comprehensive care and learning outcomes (Hong, 2023). During the COVID-19 pandemic, which saw a significant number of deaths, the incorporation of distance education could lead to more suitable educational methods (Liu and He, 2023)

This study aims to apply Advance Care Planning (ACP) and Advance Directives (AD) as key mechanisms to uphold patients’ rights to self-determination in Taiwan’s super-aged society. However, it faces challenges due to the low signing rate of Advance Directives (AD), which complicates the promotion of the Patient Autonomy Act (Zhang Zhaotian, 2024), and strategies are needed. Additionally, past research on medical autonomy and the concept of a good death has largely focused on terminal cancer patients and their families, while discussions on good death issues for healthy older adults have been relatively scarce (Li Peifang, 2020; Xu Yixiong, 2020). The research design targets older learners with clear awareness and decision-making capabilities (Liao Yingjin et al., 2022). It implements multimedia-based life education teaching to educate older learners about concepts of a good death and their rights, aiming to enhance their understanding of legal and healthy aging. This model exposes learners to knowledge related to end-of-life issues, promoting their understanding of a good death and improving their autonomy and decision-making abilities in facing the end of life (Zhang Zhaotian, 2024). The study also analyzes the impact of the intervention on the hypotheses, aiming to fill the gap in the literature regarding the concept of a good death among healthy older adults. Additionally, it seeks to provide a more humanized care system for older learners’ medical autonomy and dignity at the end of life, and to serve as a reference and suggestion for establishing comprehensive life education.

## Materials and methods

### Design and setting

This study employs a quasi-experimental design based on a literature review, analyzing the impact of multimedia life education interventions on older learners in southern Taiwan (Tainan City, Kaohsiung City, and Pingtung County) from four dimensions: “a peaceful death,” “death awareness,” “informed consent,” and “medical autonomy.” The research examines the perceived effectiveness of the concept of a good death and its influence on decision-making capabilities, aiming to promote a humane care environment and enhance the quality of life and autonomy of older adults. The findings are expected to provide important references for life education and offer recommendations regarding end-of-life and good death issues that older individuals will face.This study proposes four hypotheses:

(1).The experimental group will have significantly higher scores in the dimension of medical autonomy compared to the control group.

(2).The experimental group will have significantly higher scores in the dimension of peaceful passing compared to the control group.

(3).The experimental group will have significantly higher scores in the dimension of death awareness compared to the control group.

(4).The experimental group will have significantly higher scores in the dimension of informed consent compared to the control group.

### Sampling strategy and data collection

This study employs a cross-sectional survey method, using a questionnaire as the data collection tool. The concept of a good death is assessed using the short version of the Miyashita Good Death Questionnaire, along with additional items related to medical autonomy for older adults. Based on prior research titled “An Initial Exploration of the Practice of the Patient Autonomy Act,” data analysis is conducted using SPSS 22.0 and AMOS 22.0, confirming that the model for the four dimensions meets the requirements of positive definiteness. The questionnaire consists of three parts: basic information, which surveys participants’ gender, age, marital status, etc.; the good death concept scale, which utilizes the Chinese version of the Miyashita short questionnaire; and the medical autonomy scale, which is a self-developed questionnaire assessing older adults’ awareness of medical autonomy. A seven-point Likert scale is used. The research results indicate that the overall Cronbach’s α value is 0.936, demonstrating good reliability. This study recruited learners aged 55 and older from lifelong learning institutions in southern Taiwan, ensuring that they had clear cognition and showed no signs of dementia.

The data collection period was from September 2021 to December 2021. Participants were randomly assigned to the experimental group (216 individuals) and the control group (136 individuals). Questionnaires were distributed to qualifying elderly participants only after obtaining informed consent from each individual, followed by a pre-test. During the study, both groups received the same teaching content; the control group received traditional verbal instruction, while the experimental group was provided with educational materials through multimedia teaching interventions. The videos used were approved by governmental health authorities and produced by the Taiwan Hospice Foundation. These videos were presented in three commonly used languages in southern Taiwan (Mandarin, Taiwanese, and sign language) to accommodate the diverse cultural learning needs and cognitive abilities of the elderly. After completing the research intervention, questionnaires were distributed to qualifying elderly participants again, following the same informed consent process, for a post-test. The research process is illustrated in Figure 1. A total of 346 valid questionnaires were collected (214 from the experimental group and 132 from the control group; 6 participants withdrew due to unwillingness to discuss end-of-life issues), resulting in a response rate of 97.8%.

**Figure 1.**
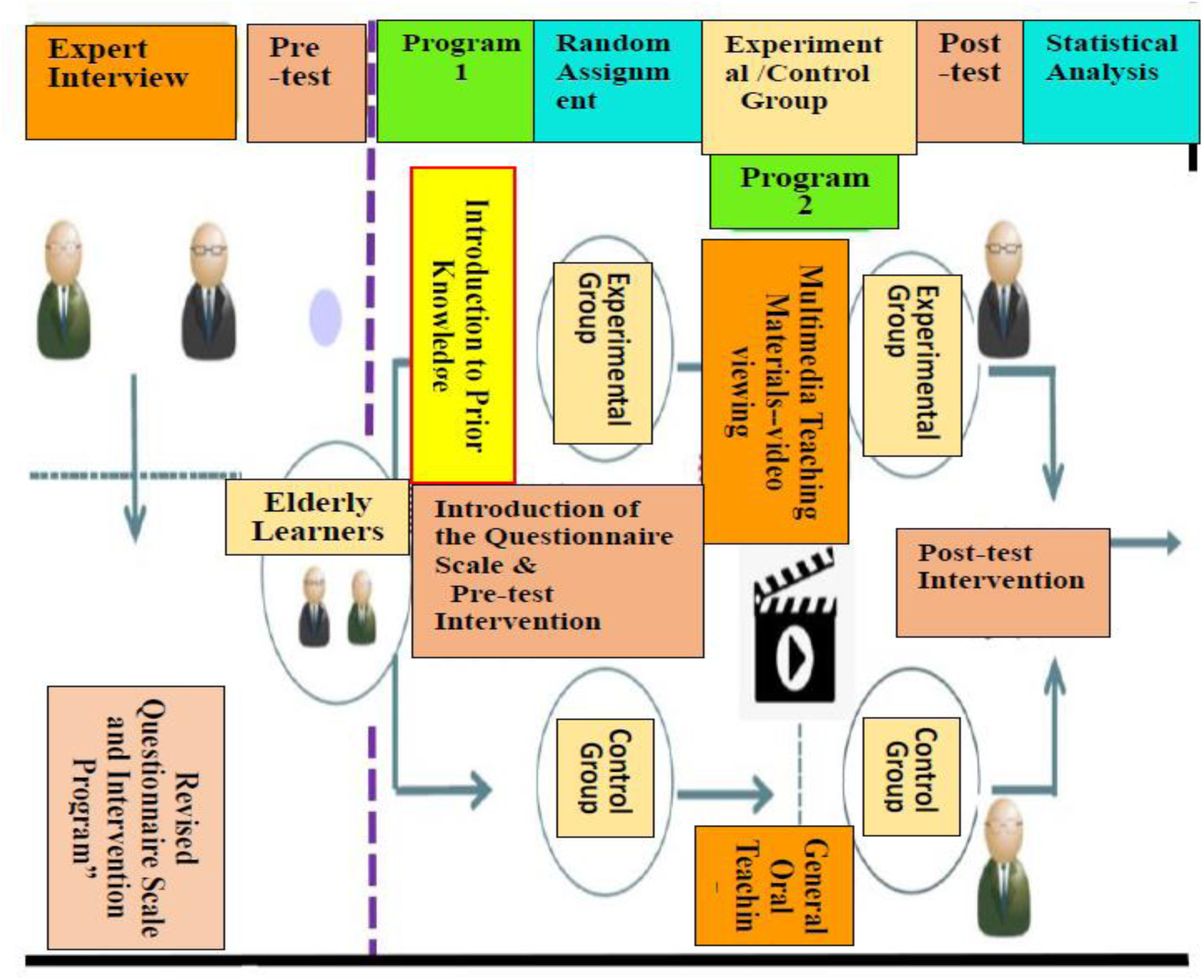
Experimental Research Diagram.

### Data analysis

Data processing in this study utilized repeated measures t-tests and analysis of covariance, with data analysis conducted using SPSS 22.0. The research tools underwent item analysis, exploratory factor analysis, reliability analysis, and structural equation modeling validation.

### Ethical considerations

This study underwent ethical review by the Institutional Review Board of Antai Medical Care Corporation Antai Hospital in Pingtung County, Taiwan, and was approved (Approval Number: 21-061-B; Approval Date: May 4, 2021; Valid Until: May 3, 2022).Before filling out the questionnaires, the researchers contacted each older participant and provided them with an informed consent form. This form included information on the purpose of the study, its content, points of attention, participants’ rights and privacy, as well as the affiliation of the researchers. Participants who agreed to participate confirmed that they understood the relevant information and voluntarily chose to take part. The questionnaires were distributed to eligible older participants only after obtaining their consent. For participants who were unable to read the consent form or had reading difficulties, the researchers provided assistance by reading the questionnaire aloud and obtained consent from the older participants prior to administering both the pre-test and post-test questionnaires. All questionnaires will be completed anonymously, and upon collection, they will be assigned identification numbers. Strict confidentiality measures will be implemented: paper data will be stored in a locked filing cabinet for two years, while electronic data will be password-protected to ensure the security of participant information. Through a series of ethical guidelines and protective measures, we aim to create a safe and respectful environment that promotes and ensures that older adults receive the necessary rights and support at every stage of participation.

## Results and discussion

The gender distribution of the study participants was predominantly female; however, apart from gender, health status, and education level, these three factors did not show significant effects on the concept of a good death. Additionally, univariate and dual covariance analyses confirmed a correlation between age and the good death concept scale, indicating significant differences in the age variable regarding peaceful death and death cognition. For example, items such as “not being treated as an object or child,” “feeling that one’s life is complete,” and “maintaining one’s role in family or professional settings” showed these differences. Furthermore, the results revealed a significant association between marital status (married vs. unmarried) and the dimension of peaceful death. However, gender, health status, and education level did not significantly influence the concept of a good death, and the impact of religious belief and occupational distribution on the concept was also rather limited. This may be related to the participants being older (the overall average age of the subjects was 73.45 years) and having a more consistent religious belief system (34.4% Buddhist and 45.6% Shinto). Additionally, items such as “being able to stay in the preferred location,” “the right to refuse treatment,” and “the right to choose treatment” showed that the t-values of the experimental group had p-values less than 0.001, indicating a significant enhancement in participants’ autonomy in decision-making after the intervention (see Table 1).

**Table 1:** The Differential Effects of Remote Education Intervention on End-of-Life Perspectives of Learners in Senior Centers with Different Background Variables.

| Two-Way Analysis of Covariance for Group Differences on Dimensions and Item Content. | Marital Status | Physical Health | Gender | Education | Age |
| --- | --- | --- | --- | --- | --- |
| Peaceful Passing | O (.049) | X (.320) | X (.468) | X (.128) | O (.003) |
| Informed Consent | X (.858) | X (.805) | X (.691) | X (.420) | X (.086) |
| Death Awareness | X (.321) | X (.406) | X (.933) | X (.849) | O (.002) |
| Medical Autonomy | X (.656) | X (.347) | X (.280) | X (.541) | X (.766) |

O indicates significant; X indicates not significant. \*Age covariate: pre-test scores and age, independent variable: group, dependent variable: post-test scores.

The results of this study indicate that women have a greater awareness of the importance of “being free from pain and physical discomfort” in the process of a good death, reflecting gender-related cognitive and emotional differences in end-of-life care.

The research analysis and hypothesis testing showed that the experimental group scored significantly higher than the control group in various dimensions of peaceful death and death cognition. However, in the dimensions of informed consent and medical autonomy, the experimental group did not demonstrate a significant advantage over the control group. Comparing the pre- and post-test scores of multimedia teaching and traditional teaching revealed that both intervention methods significantly improved aspects of peaceful death, informed consent, death cognition, and medical autonomy. This highlights the substantial learning needs of older adults regarding the concept of a good death.

It is worth noting that for the item “resisting illness at the last moment,” the experimental group’s t-value and p-value were both less than 0.001, indicating a significant attitude change after the educational intervention. They understood that continuing to fight against death would lead to more suffering and ineffective medical treatment. For items such as “being able to stay in a place they love,” “the right to refuse treatment,” and “the right to choose treatment,” the experimental group’s t-values also showed p-values less than 0.001, demonstrating a significant improvement in participants’ decision-making autonomy (see Table 2).In certain themes, such as “not being a burden to family,” “I have the right to refuse treatment,” “natural death,” and “resisting illness at the last moment,” which relate to the right to refuse ineffective medical treatment, as well as items like “being able to foresee future conditions,” “not realizing that I am approaching death,” “I have the right to fully understand my condition,” “I have the right to consent to treatment,” and “I have the right to choose treatment,” which are related to informed medical consent and the shared decision-making process, this study observed significant changes.The descriptive analysis of the end-of-life questionnaire indicated that the experimental group scored highest on the item “natural death,” reflecting a positive understanding of a peaceful death. In contrast, the control group scored highest on items related to “medical autonomy,” suggesting that promoting awareness of medical autonomy can be effectively achieved with positive interventions. It appears that the challenges in promoting advance directives (A.D.) may be attributed to insufficient advocacy!

**Table 2.**
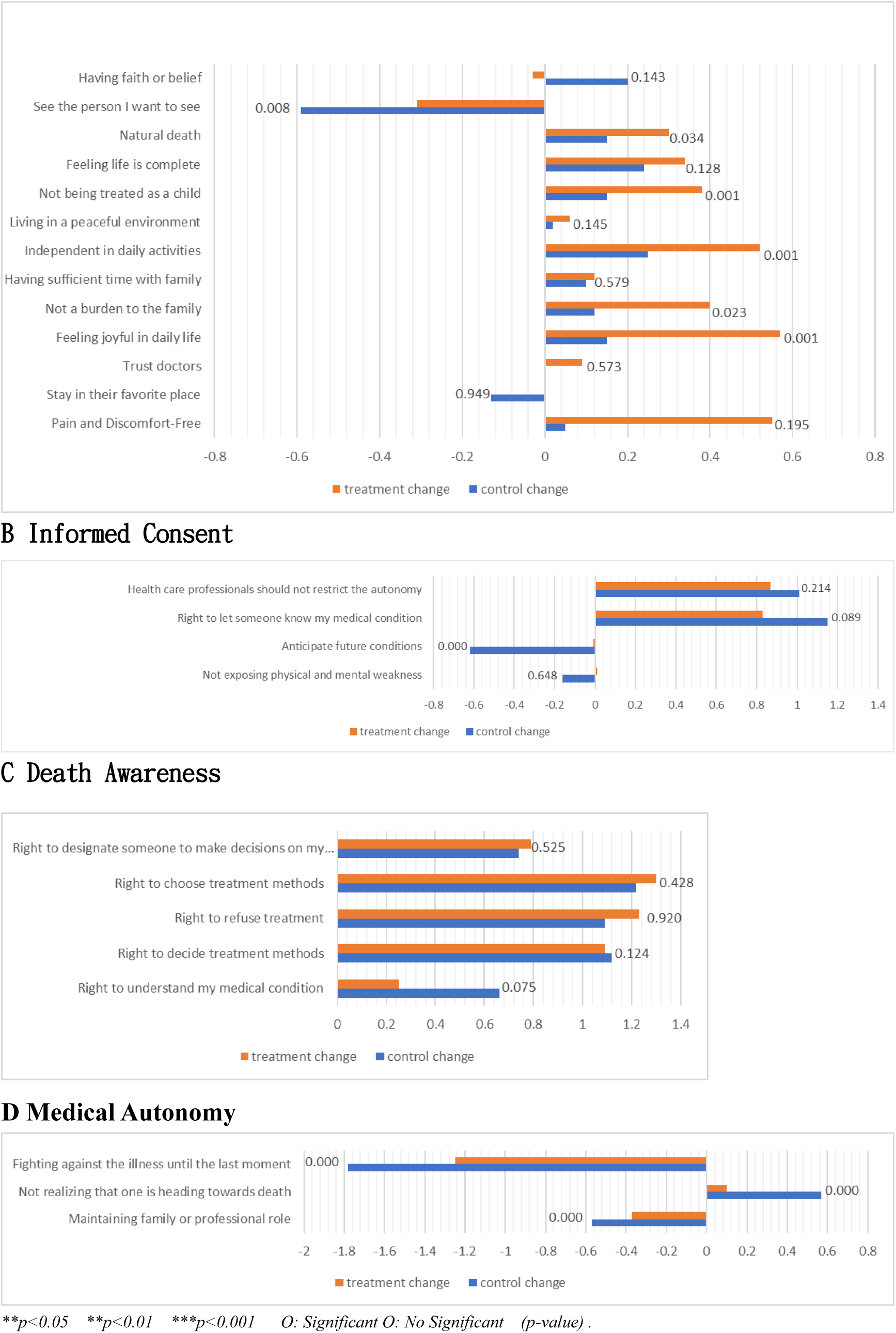
The Effects of Remote Education Intervention Program on End-of-Life Concepts.

Furthermore, the data indicated that older participants significantly improved their scores in areas such as “natural death,” “enjoyment in daily life,” and “medical autonomy” through the program intervention, highlighting a substantial need for end-of-life education among older adults. When examining the questionnaire regarding “maintaining roles in family or professional environments,” it was noted that social changes profoundly affect the psychological and emotional states of older adults, due to traditional societal expectations of their roles in the past. These rigid roles may impose unnecessary stress and anxiety on older individuals (see Table 2).

### Research Contributions & Discussion

The relevant scales used in this study have mostly been applied in the context of cancer or terminal palliative care patients and healthcare professionals. In contrast, the subjects of this study were older participants not under the stress of illness. The research explores the application of multimedia life education interventions, which broadens and enhances the flexibility of the applicability of these scales, effectively reflecting the needs of different groups and improving doctor-patient interactions. This study validates the literature and contributes in the following ways:

### population demographic variables

Gender often influences the approach to life education, such as: “Choosing different content based on the focus of different topics” (Wang Lijing, 2025; Zhu Meizhen et al., 2015); “Women tend to prefer collaborative learning and discussion, while men are more inclined towards competitive or task-oriented methods” (Zheng Wen et al., 2022); “Women often excel in emotional expression and empathy, while men require more guidance to understand and express emotions” (Jiang Wenci, 2018); “Gender expression varies across different cultures” (Zhang Fangquan, 2024; Guo Fuzi et al., 2012), and “is influenced by social expectations and pressures” (Hong Peilun et al., 2025). Therefore, in future teaching models focused on life education and death cognition for older adults, it may be necessary to consider adjustments based on gender differences to better meet the diverse needs of learners and promote their comprehensive development.

Age also has a significant impact on the approach to life education. For example: “Different age groups possess different knowledge and experiences” (Chen Yujing et al., 2025); “With increasing age, cognitive abilities and abstract thinking skills often improve” (Chen Yiling et al., 2018); “The emotional needs of different age groups also vary” (Lin Jiajing et al., 2018); “Learning styles and preferences change with age” (Lin Yijun, 2023); and “The needs formed by different socio-cultural backgrounds also differ” (Zeng Beilu, 2021). Therefore, life education should focus on the influence of age on the concept of a “good death,” designing customized curricula and integrating emerging technologies to meet the needs of learners. This approach will promote learners’ understanding and acceptance of the end of life, enhance self-care abilities, and improve doctor-patient relationships.

Over the past 30 years, the proportion of individuals in Taiwan who remain lifelong single has gradually increased. Although the impact of individual socioeconomic conditions on being single varies by gender, the proportion of single men and women living with their parents has also risen. However, single women often bear a greater burden of caring for aging parents. Therefore, life education should also recognize the necessity and uniqueness of addressing needs related to marital status (Yang Jingli et al., 2023).

The factor of education level has a significant impact on the utilization of healthcare resources and health behaviors. In terms of teaching models for life education, for example: “Individuals with higher education levels typically have a richer knowledge base” (Lin Yijun, 2021); “Critical thinking and analytical skills are enhanced through higher education” (Chen Guanshao, 2021); “Individuals with lower education levels may rely more on teacher guidance and direct instruction” (Yang Shiyu et al., 2025); “Those with higher education often have more social experiences and diverse perspectives” (Chen Guanghui, 2019); and “There are also differences in values and emotional expression based on education levels” (Lin Naihui et al., 2021). Therefore, it is necessary to consider whether adjustments to the teaching model of life education should be made based on education levels to better meet the needs of learners.

Incorporating a religious perspective can provide important support for advance care planning and end-of-life care. The influence of religious beliefs on the understanding of life and death aids in facilitating effective communication between patients and medical teams, reducing conflicts and promoting consensus in medical decision-making. Furthermore, religious beliefs also impact the teaching of life education: “They influence the understanding of issues related to life, death, ethics, and morality” (Lin Mengqian, 2024); “The selection of teaching materials and case studies varies based on religion” (Chen Xinjue, 2021); “Discussions on life education may hold different viewpoints and perspectives” (Chen Meini, 2023; Fan Chongguang, 2016); “Religious beliefs can promote reflection on significant issues in life, such as suffering and loss” (Lin Zimin, 2025); “Encourages students to be more willing to participate in community activities” (Fan Ganghua, 2020); “Provides a support system for facing difficulties and challenges” (Jian Jingwei, 2014); “Enhances inclusivity and empathy” (Zhang Mingci, 2022); “Influences the understanding and handling of ethical and moral issues” (Chen Yijun, 2022); “Identifies and ritualizes different life stages (such as birth, growth, and death)” (You Huizhen, 2011); and “Provides emotional support when facing life’s challenges” (Fan Ganghua, 2020).

Therefore, applying the role of religious beliefs in end-of-life care within life education and encouraging healthcare professionals to respect and integrate patients’ religious backgrounds can enhance the quality of medical care and increase patient satisfaction, thereby achieving more humane medical services. Designing corresponding life education curricula based on faith backgrounds will facilitate holistic and comprehensive growth.

### Implications in research and practice

In residents of southern Taiwan, personalities are generally more introverted, and they are less adept at expressing their emotions. Consequently, their willingness to express views on issues related to life and death is also lower (Zhang Zhaotian, 2024).This characteristic is one of the reasons why the author chose elderly learners from this region as the subjects of this study.

However, learners’ self-expression abilities may influence the teaching models of life education, subsequently affecting their ability to “set independent thinking and diverse evaluation perspectives as learning goals, thus changing their lifelong learning capacity and resource application” (Huang Qingmei, 2022); “engage in creative thinking on life education topics, which impacts their complex analytical abilities regarding the meaning of life and interpersonal relationships” (Zheng Yaotiao, 2023); “analyze practical situations in ethical dilemmas more effectively” (Cai Changying, 2020); “understand their own beliefs and values more deeply, thereby improving self-expression and comprehension skills” (Zhan Fuxuan et al., 2024); “encourage reflection on how to responsibly face social challenges, promote understanding and respect for diverse cultures, and build healthier interpersonal relationships” (Li Shengwei et al., 2024); “increase interactions between teachers and students, thereby enhancing their motivation to learn and promoting collective thinking and innovative collaborative learning” (Chen Meiling, 2022); as well as “guide students to engage in regular self-reflection to cultivate critical thinking skills and assess their own expressive abilities and growth” (Chen Guanshao, 2021). By effectively enhancing students’ self-expression abilities, not only is the learning effect of life education strengthened (Zhang Haozhi et al., 2020), but a solid foundation is also laid for future development.Research indicates that intervention measures successfully changed elderly learners’ awareness of health rights and responsibilities, enhancing their legal competence regarding health autonomy (Chou Tzuyu, 2021). This demonstrates that through appropriate educational interventions, it is possible to improve learners’ willingness to express themselves autonomously. They can actively demonstrate confidence in their health decisions regarding medical choices and the right to refuse, thereby strengthening the doctor-patient relationship and improving the overall healthcare experience (Lin Huifang, 2022). This enhancement in self-expression willingness can increase their confidence in actively expressing medical choices, exercising the right to refuse, and making health decisions. Clearly, designing more customized courses for different social groups not only enhances elderly learners’ understanding of medical autonomy but also helps reduce fear of death, improve quality of life, and promote mental health and coping abilities for illnesses. This, in turn, fosters a sense of dignity and value at all stages of life, leading to a greater understanding and acceptance of a good death.

The behavior of “not exposing physical and mental vulnerabilities in front of family” may be related to the super-aged society, combined with trends of declining birth rates and socioeconomic conditions. Elderly individuals may choose to hide their weaknesses out of fear of becoming a burden, which not only affects their mental health but may also delay access to medical support, leaving them to face loneliness and the stress of suffering. Therefore, the practice of life education should strengthen open communication within families and encourage elderly individuals to express their needs and feelings as much as possible. Additionally, the item “being able to stay in a favorite place” refers to the choice of the place of death. Research has found that this choice is closely related to individuals’ quality of life and dignity. According to studies, “dying at home” is considered a core attribute of a good death, reflecting the Chinese cultural concept of “returning to one’s roots.” It aligns with the wishes of individuals in Eastern societies who hope to peacefully depart in their homes or familiar environments, which is an important consideration for a good death. This perspective is particularly emphasized among elderly learners in southern Taiwan (Liu Qianwei, 2019). With advancements in technology, while it is not possible to treat all critically ill patients, the demand for medical care still exists (Su Jiehan, 2022). Many patients return to emergency care hospitals in their last month of life, which not only diminishes the quality of end-of-life care but also increases medical costs (Xu Shaoen, 2023; Lage et al., 2018). Therefore, establishing a humane end-of-life care system that respects patients’ choices and needs is particularly important. Furthermore, research indicates that men are more likely than women to choose to die at home (Lin Xinyi et al., 2025; Choi K.S. et al., 2005; Choi J.E. et al., 2010). Consequently, when designing life education courses, it is essential to consider the influence of gender on the choice of end-of-life location.

Due to emotional factors, many family members choose to prolong cardiopulmonary resuscitation or ineffective medical treatment in order to allow loved ones to have one last meeting with the dying individual. This often results in unnecessary suffering for the patient. Therefore, when exploring the questionnaire regarding “people they hope to see,” it was found that enhancing life education can improve families’ understanding of the end-of-life stage. This understanding is crucial for helping them face end-of-life decisions more rationally and reducing unnecessary medical interventions.

On the other hand, it was found that elderly individuals who received life education showed significant improvements in mental health, enabling them to adapt flexibly to their roles, establish good communication and support systems, and reduce levels of anxiety and stress. Research indicated that when exploring the item “fighting illness until the last moment,” many elderly people, due to traditional beliefs, tend to overlook the importance of quality of life and choose to undergo prolonged cardiopulmonary resuscitation or ineffective medical treatments. However, through interventions in life education, the participants’ perspectives underwent significant changes. Although the research showed that some post-test scores were lower than pre-test scores, this reflects the elderly’s rethinking of the end of life. Participants began to understand that prolonging unnecessary medical interventions does not equate to respecting life; rather, it may increase suffering. They gradually accepted the concept of a good death, choosing to receive dignified care in their final days. In other words, life education can enhance the potential of elderly individuals to comprehend the notion of a peaceful death and the reality of dying, although education on medical autonomy still needs to be strengthened. Ultimately, effective teaching interventions can help the elderly better understand and confront end-of-life choices, thereby improving their quality of life and mental well-being. Through these findings, this study promotes the development of life education, successfully enhancing the understanding of elderly individuals facing end-of-life issues and facilitating their awareness and acceptance of death, ultimately providing them with more comfort and support. For future improvements in scale design, it is recommended to convert such items into reverse questions to more effectively assess participants’ attitude changes and strengthen their recognition of the concept of a good death.

Through these findings, this study promotes the development of life education, successfully enhancing the understanding of elderly individuals facing end-of-life issues, and fostering their awareness and acceptance of death, thus providing them with more comfort and support. For future improvements in scale design, it is recommended that items exploring the desire to prolong end-of-life survival time be formulated as reverse questions to more effectively assess changes in participants’ attitudes and reinforce their recognition of the concept of a good death. This new understanding of rights and obligations regarding the use of social healthcare resources also emphasizes the cultivation of legal literacy and the ability for autonomous medical decision-making within the practice of life education (Chou Tzuyu, 2021). Through such educational models, learners can effectively enhance their understanding of the meaning of life, especially in crisis situations at the end of life, where it is crucial to balance the rights and responsibilities between patients and caregivers, prompting a reassessment of personal values and critical reflection. Through rational dialogue, individuals can form new understandings of life’s meaning and values (He Yiwen, 2022).It is also hoped that through education and advocacy, understanding of end-of-life choices for the elderly within families and society can be enhanced, which will help improve the quality of life and care at the end of life. This includes understanding feasible options for home hospice care and the necessary support systems, as well as effectively communicating with the medical team to ensure that patients’ needs are adequately addressed.

### Doctor-patient communication

In certain items of this study, such as “not becoming a burden to my family,” “I have the right to refuse treatment,” “natural death,” and “resisting illness at the last moment,” these items relate to the right to refuse ineffective medical treatment. Additionally, items like “being able to foresee future conditions,” “not realizing I am approaching death,” “I have the right to fully understand my condition,” “I have the right to agree to treatment,” and “I have the right to choose treatment” are related to informed medical consent and the shared decision-making process. If these can be integrated with communication skills in doctor-patient interactions (Zhang Qingyan, 2022), and combined with knowledge from fields beyond biomedicine—such as psychology, sociology, and ethics—to comprehensively enhance communication skills within life education, it would further cultivate healthcare professionals’ humanistic care and improve the support and understanding for elderly individuals facing end-of-life issues. This would also provide healthcare personnel who receive death education training with a high level of self-awareness for self-protection and professional development, as well as a well-rounded workplace education and training environment (Huang Yichen, 2023).

In clinical practice, a lack of interaction is most likely to lead to blind spots and disputes between doctors and patients (Huang Guankai, Zheng Bowen, 2018). Literature indicates that multimedia teaching using technology can enable educators to effectively grasp teaching strategies while also focusing on the learning motivations of older learners. In teaching doctor-patient communication, specific course content can be selected to enhance the construction of simulated conversations and legal competence, thereby improving legal literacy and emphasizing communication skills. In multimedia teaching for the elderly, the autonomy of learners and the communication attitudes of both parties are also key factors. Future efforts should aim to enhance the understanding and acceptance of end-of-life concepts among elderly learners (Chou Tzuyu, 2021). This study successfully highlights the importance of life education, improving learners’ knowledge, skills, and attitudes in doctor-patient communication. The results also indicate that it contributes to promoting humanized medical services and improving doctor-patient relationships.

### Transformative learning

Transformative learning is particularly critical in the context of elderly education, requiring older learners to reflect and explore the value and beliefs of their own lives, thereby revising and acquiring new knowledge. As mentioned earlier, it was found that elderly individuals who received life education showed significant improvements in their mental health. The research also indicates that regarding the item “to resist illness until the last moment,” participants’ perspectives underwent significant changes due to the intervention of life education. The impact of transformative learning on the teaching model of life education, as stated, is that “transformative learning in elderly education promotes a deeper understanding and transformation of their own life experiences, helping them to re-reflect and reassess their values and beliefs” (Liu Jiaqi, 2019); “it shifts the role of teachers from knowledge transmitters to facilitators and guides” (Ye Juntao, 2019); “it encourages older learners to express their feelings and viewpoints, thus promoting deeper emotional understanding and connections” (Wei Caimi, 2019); and “it enables older learners to apply the values and skills they have learned to their daily lives, becoming lifelong learners” (Zhang Wanzhen, 2023). Transformative learning offers a more dynamic and reflective teaching model for life education in older learners, facilitating personal engagement through subjective experiences and emotions, deepening their understanding of the meaning of life and end-of-life concepts, enhancing their coping abilities, and promoting healthy behaviors and effective utilization of medical resources.

### The Impact of Multimedia Teaching

When implementing multimedia teaching, sufficient interaction is expected to achieve better results. However, this study indicates that the combination of traditional PPT and multimedia has limited effectiveness in enhancing learning outcomes. A comparison between Procedure 1 (”My Body, My Choice”) and Procedure 2 (using public videos to discuss informed consent) shows that Procedure 1 had a significant intervention effect. This highlights the importance of establishing learners’ prior knowledge and promoting transformative learning motivation in the multimedia teaching of life education. Multimedia teaching in life education “can incorporate images, videos, and audio to provide a richer learning experience” (Huang Mengming, 2020); “can create simulated scenarios, such as life and death and ethical dilemmas, to explore important issues of life” (Lin Shangjie, Wang Jialing, 2020); “can introduce interactive elements to promote thinking based on learning needs and progress” (Hu Jiawen, 2022); “can easily integrate knowledge from different disciplines” (Hong Yongzhou, 2022); “using elements such as documentaries and real-life stories can evoke emotions and foster deeper thinking and reflection” (Lin Xiuyi, Chen Yujing, 2024); “with appropriate assessment tools, can provide immediate assessment of progress and feedback on understanding” (Chen Zhenjun, Yang Shuqing, 2022); “can introduce perspectives from different cultures and religions”; and “can easily share learning outcomes and viewpoints, facilitating collaborative learning” (Zhang Haozhi et al., 2020). Therefore, multimedia teaching offers a more flexible, engaging, and participatory teaching model for life education. It not only enhances students’ learning motivation but also strengthens their understanding and critical thinking skills, helping them gain a more comprehensive understanding of the meaning and value of life.

### The Impact of Prior Knowledge

If appropriate foundational knowledge on issues of life and death is provided, such as enhancing awareness of death and exploring the meaning of life, it can “give elderly learners greater intrinsic motivation, maintain a high level of interest and drive in life education, encourage them to explore more deeply, integrate into the course content more quickly, and establish stronger connections in new learning; they can also engage in deeper analysis and pose more profound discussion questions” (Zhang Jingyuan, Hu Minhwa, 2015); “they are able to develop clearer personal ethical views and value systems, leading to more mature thinking” (Lai Zhizhan, 2024); “they resonate with the content, are able to share personal experiences, and are more willing to express their viewpoints; participants often engage in dialogue with greater confidence, clearly articulate their opinions, understand others’ emotional perspectives, and enhance communication among peers” (Li Peiyi, 2004). “The curriculum can include more challenging case studies and in-depth discussions; learners can more effectively apply what they have learned in simulated scenarios or role-playing, enhancing their practical coping skills” (Cai Miaojuan, 2013); “learners will have stronger self-reflection abilities and critical reflection on real-life situations”; “they can better respect diverse cultural perspectives and show increased interest in professions related to life and death (such as healthcare and psychological counseling), demonstrating a sense of social responsibility” (Sun Zhichen, 2024); “they can view death more rationally, reduce the fear of death, and enhance psychological resilience” (Cai Miaojuan, 2013). These foundational knowledge components of life and death education can appropriately address the aforementioned considerations regarding the relative costs of prolonging life and the dignity of life quality through teaching strategies. This allows learners to engage in deeper thinking and emotional resonance within the life education model.

### Research Strengths

This study aims to explore the impact of life education on elderly learners regarding medical autonomy and end-of-life concepts. The research subjects are a group of elderly learners in southern Taiwan who possess independent learning and thinking abilities and are not facing the pressures of life and death, yet are reflecting on the meaning of life and medical decision-making (Li Meifang, 2021). Therefore, it excludes similar subjects from past research, which mostly focused on terminal cancer patients, their families, and healthcare professionals. The research results indicate that after participating in life education with multimedia resources, elderly learners significantly improved their confidence and sense of involvement in medical choices, making them more proactive and independent in medical decision-making. This enhancement promotes a greater awareness of personal health and quality of life, as well as a better understanding of medical autonomy and end-of-life concepts (Chou Tzuyu, 2021). This transformation not only improves the psychological health of the learners but also provides a clearer communication foundation for families and healthcare teams (Yan Peihan, 2024). Furthermore, the study shows that the involvement of family members can enhance the learning motivation and confidence of elderly individuals. If multimedia resources or distance learning platforms that overcome geographical limitations can be utilized to facilitate intergenerational education and encourage family members to participate in providing psychological support and communication with the elderly (Chen Yujing et al., 2025), it would be highly meaningful for the elderly to reassess the value of life and advocate for humane and dignified care approaches at the end of life.

The new questionnaire designed for this study provides a comprehensive assessment to explore the cognitive attitudes and specific needs of elderly individuals regarding end-of-life care and medical autonomy. Through the analysis of the research data, the attitudes and behavioral patterns of elderly individuals in medical decision-making are revealed. The findings not only deepen the understanding of medical autonomy among the elderly but also enhance the emphasis on quality of life. The validity and reliability of the questionnaire have been verified, demonstrating its value in quasi-experimental research. This provides an empirical foundation and effective tools and methods for future life education programs, while also promoting the formulation of relevant policies to create a more friendly healthcare environment.

### Research limitations

Although this study designed a comprehensive questionnaire to assess the cognitive attitudes of elderly individuals regarding medical autonomy and end-of-life decision-making, it still faces some limitations. First, the respondents’ answers may be influenced by personal factors, environmental contexts, and health conditions, leading to potential data bias. Second, the lack of diversity in the sample may not fully represent the elderly population, particularly concerning cultural, economic, and educational backgrounds, which could affect the generalizability of the results. Therefore, future research should expand the sample range to enhance external validity. Subjective limitations among participants, such as some elderly individuals refusing to engage with the topic of “end-of-life” due to cultural or psychological factors, can lead to decreased participation; the researcher effect; the influence of education level and physiological conditions on questionnaire comprehension; and the accuracy of questionnaire translation, all of which may affect data validity. In summary, future studies in life education should pay attention to these subjective limitations and adopt diversified research methods to minimize the interference of cultural and language factors, thereby improving and ensuring elderly individuals’ understanding and expression regarding end-of-life care and medical autonomy.

## Conclusions

The French philosopher Voltaire once said that humans are the only beings aware of their own mortality. However, it is crucial for both families and patients to adopt a mindset that enables them to face illness and death with peace. As Taiwan moves into a super-aged society, the medical autonomy choices related to health issues among the elderly and their desire for dignified death are increasingly prominent. Therefore, it is essential to promote accurate understanding of relevant choices among the elderly and their families, assisting them in making informed health management and end-of-life decisions. This includes physical, psychological, spiritual, and social interactions, as well as the integration of medical resources to provide appropriate care, highlighting the necessity of offering dignity in end-of-life care and related life education.

This study explored the understanding of end-of-life concepts among elderly individuals in southern Taiwan. The results showed that multimedia teaching provided a flexible learning model that enhanced learners’ sense of participation. Through interactivity and visual design, combined with comprehensive foundational knowledge, it effectively helped elderly learners grasp and deepen their understanding of complex concepts such as medical autonomy and dignified death. After the intervention, learners exhibited significant changes in cognition, improving their awareness of death and the effectiveness of end-of-life concepts while reducing the acceptance of ineffective medical treatments due to fear and misunderstanding.

The newly developed end-of-life concept scale effectively enhances learners’ health literacy and legal awareness, strengthening their autonomy and decision-making abilities when facing death and medical choices. The study emphasizes the centrality of elderly individuals’ needs and the importance of autonomous choice. It also suggests encouraging family involvement to break traditional patriarchal views, fostering understanding and support among family members, ultimately enhancing quality of life and dignity.

The study highlights the importance of innovative multimedia teaching models in life education and their profound impact on promoting a person-centered care environment. It fosters consensus in the social, medical, and legal fields regarding end-of-life care and medical autonomy, helping to restore harmony and trust between patients and healthcare providers while reducing medical disputes and resource wastage. Effective teaching interventions not only change learners’ cognition but also provide references for overcoming the challenges of promoting medical autonomy. Additionally, it offers guidance and basis for future government policies concerning the well-being of the elderly, ensuring that everyone can attain the dignity and medical autonomy they deserve at the end of life.

## Data Availability

All relevant data are within the manuscript and its Supporting Information files.

## Notes

**Conflicts of Interest**: The authors declare no conflicts of interest.

### Competing Interest Statement

The authors have declared no competing interest.

### Funding Statement

The author(s) received no specific funding for this work.

### Author Declarations

This study underwent ethical review by the Institutional Review Board of Antai Medical Care Corporation Antai Hospital in Pingtung County, Taiwan, and was approved (Approval Number: 21-061-B; Approval Date: May 4, 2021; Valid Until: May 3, 2022).

### Summary of Updates

no revision has been made to this version of the manuscript

## References

1. Chang CM, Lee CL, Lin HC. Advance Directives in Older Adults. Taiwan Geriatrics & Gerontology. 2016;11(2):89–104.

2. Chang, Chao-Tien. Advance Autonomy, Dynamic Autonomy, Relational Autonomy: Reflecting on the Legal and Ethical Issues of Advance Medical Decisions Under the Patient Autonomy Act. Taiwan Law Review, 2024, No. 345, p. 119.

3. Chang, Chao-Tien. Advance Autonomy, Fluid Autonomy, and Relational Autonomy: Reflecting on the Legal and Ethical Issues of Advance Medical Decisions Under the Patient Autonomy Act. Taiwan Law Review, 2024, No. 345, p. 119.

4. Chang, Ching-Yen. A Study on Euthanasia and Assisted Suicide from the Perspective of the Doctor-Patient Relationship. Master’s Thesis, National Taiwan University, 2022.

5. Chang, Fang-Chuan. Exploring National Development from the Perspective of Gender Equality. School Administrator, 2024, Vol. 149, p. 17. DOI: 10.6423/HHHC.202401_(149).0002.

6. Chang, Hao-Chih, and Chen, Ruo-Fan. Autonomous Learning Weeks and the Transformation of Teacher Roles: Educational Innovations and Practices under the New 16-Week Semester System. Taiwan Education Review Monthly, 2025, 14(5), pp. 43–52.

7. Chang, Jing-Yuan, and Hu, Min-Hua. The Connotation and Practice of Life Education. National Academy for Educational Research Educational Pulse Electronic Journal, September 2015, Issue 3.

8. Chang, Ming-Tzu. A Study of the “God” and the Life Care Contained in “Conversations with God.” Master’s Thesis, Graduate Institute of Religious Studies, Nanhua University, 2022.

9. Chang, Wan-Chen. A Study on the Effectiveness and Transformative Learning of College Students Participating in Service Learning for Community-Dwelling Elderly with Dementia. Journal of Educational Science Research, 2023, Vol. 68, No. 2, p. 235.

10. Chen, Cheng-Chun, & Yang, Shu-Ching. A Review and Outlook on Technology-Enhanced Language Teaching and Learning. Curriculum and Teaching Quarterly, 2022, 25(1), pp. 135–172.

11. Chen, Guang-Hui. Rethinking Educational Attainment: A Longitudinal Study on College Students’ Political Knowledge and Attitudes. Survey Research: Methods and Applications, October 2019, No. 43, pp. 45–88.

12. Chen, Hsin-Chueh. Alternate title: Exploring Diverse Views of Taiwanese Christians on Evolution and Teaching Evolution from the Perspectives of Worldview and Social Constructivism. National Taiwan Normal University (Taiwan), ProQuest Dissertations & Theses, 2021.

13. Chen, Kuan-Shao. Alternate title: A Study of Controversial Issue Courses Based on Critical Thinking Teaching Method: An Action Research of Civic and Society in Senior High School. National Taiwan Normal University (Taiwan), ProQuest Dissertations & Theses, 2021.

14. Chen, Mei-Ling. A Study on Enhancing English Learning Effectiveness through Situational Teaching and Cooperative Learning: English 2. Ministry of Education Teaching Practice Research Project Results Report, PGE1101010, September 2022.

15. Chen, Mei-Ni. Challenges and Opportunities for Catholic High Schools under the Crisis of Declining Birth Rates. Taiwan Education Review Monthly, 2023, 12(9), pp. 115–119.

16. Chen, Shih-Pei, Tseng, Fei-Lin, Huang, Kun-Hsiang, Shih, Jun-Ying, Chou, Man-Chun, & Jiang, Ming-Chu (2019). Respecting the choice of life: A brief discussion on the Patient Autonomy Act. Chang Gung Nursing, 30(4), 466–473.

17. Chen, Yi-Chun. A Research on Contemporary Neo-Confucian Zeng, Zhao-Xu’s Confucius Religiosity. Master’s Thesis, Department of Life and Death Studies, 2022.

18. Chen, Yi-Ling, Wei, Hui-Chuan, and Huang, Ching-Yun. A Study on Cognitive Function and Physical Activity in Older Adults. National Chiayi University Journal of Sports, Health and Leisure, 17(2), pp. 74–88.

19. Chen, Yu-Jing, Chen, Dai-Fen, & Chen, Jing-Yang. A Study on the Development and Strategies for Promoting Intergenerational Learning in University of the Third Age. Educational Policy Forum, 2025, Vol. 28, Issue 1, p. 95.

20. Cheng, Wen, Hsia, Pei-Chen, Chang, Ya-Feng, Wu, Hui-Ru, and Hong, Chia-Hsin. A Study on the Effects of Cooperative Learning and Narrative Teaching on Learning Motivation in Chinese Language among Students of Different Genders and Academic Achievements. Positive Psychology: Counseling and Education, No. 1, June 2022, pp. 135–161.

21. Cheng, Y. T. (2023). Deconstruction and Reconstruction: Exploring the Learning Experience of Elderly Participants in Life Story Picture Book Courses. Journal of Welfare Technology and Service Management, 11(3).

22. Chiang MC, Chou MC, Shih CY, Huang KS, Tseng FL, Chen SP. Respect for Life - Brief Introduction of Patient Right to Autonomy. Chang Gung Nursing. 2019;30(4):466–473.

23. Chien, Ching-Wei. The Impact of Coping Styles, Self-Efficacy, and Social Support on Religious Coping Under High Stress. Master’s Thesis, National Taiwan University, 2014.

24. Chien-Wei LIU, Chich-Hsiu HUNG, Wan-Ping YANG, A Challenge for Home-Based Hospice Care: Good Death at Home, The Journal of Nursing; 66, 6: 74–81 ‧

25. Choi JE, Miyashita M, Hirai K, Sato K, Morita T, Tsuneto S, Shima Y. Preference of place for end-of-life cancer care and death among bereaved Japanese families who experienced home hospice care and death of a loved one. Support Care Caner. 2010; 18:1445–1453.

26. Choi KS, Chae YM, Lee CG, Lee SW, Heo DS, Yun, YH. Factors influencing preference for place of terminal care and of death among cancer patients and their families in Korea. Support Care Cancer.2005;13: 565–572.

27. Chou TY, Wang CY, Wu ML. A Preliminary Study on Concepts of a Good Death and Patient Autonomy Act Practice. 2021;9(3):314–332.

28. Chou, Tzu-Yu. An Initial Exploration of the Practice of the Patient Autonomy Law: A Study on End-of-Life Concepts Among Seniors in Southern Taiwan. Journal of Welfare Technology and Service Management, 9(3), 2021.

29. Fan, Chong-Guang. A Study on the Inclusion of Religious Hospice Care in the Life Education Course of University Students: A Case Study Comparing Buddhist and Catholic Approaches. Master’s Thesis, Graduate Institute of Religious Studies, Nanhua University, 2016.

30. Fan, Kang-Hua. The Overlooked Positive Psychological Functions of Religion: The Impact of Psychological Distress on the Relationship Between Happiness and Religious Participation in Young Adults. Chinese Journal of Mental Health, vol. 33, no. 2, 2020, pp. 119–142.

31. Fang CK, Pi SH, Chang SY, Chen SJ, Chiu SC. Opinions and Attitudes of Patients’ Autonomy Act among the Medical Staff in Different Branches of a General Hospital. Taiwan Journal of Hospice Palliative Care. 2018;23(1):1–17.

32. Hattori K, McCubbi MA, Ishida DN. Concept analysis of good death in the Japanese community. Journal of Nursing Scholarship. 2006;38(2): 165–170.

33. He, Yi-Wen (2022). A Study on the Development of Cross-Cultural Communication Skills for International Students in Higher Education. Doctoral Dissertation, Graduate Institute of Curriculum and Instruction, National Taiwan Normal University.

34. Hirai K, Miyashita M, Morita T, Sanjo M, Uchitomi Y. Good death in Japanese cancer care: A qualitative study. Journal of Pain and Symptom Management. 2006;31(2): 140–147.

35. Hsu HL. The Experimental Construction of Philosophical Counseling and Spiritual Caring Model Based on the Gandavyūhasūtra. Universitas: Monthly Review of Philosophy and Culture. 2020;47(4):53–71.

36. Hsu, Ho-Ling (2020). Constructing a philosophical counseling and spiritual care model for life situations based on the Avatamsaka Sutra. Philosophy and Culture, 47(4), 53–71.

37. Hsu, Shaw-En (2023). Exploring the Facilitators and Barriers of Advance Care Planning Counseling for Elderly Individuals with Chronic Diseases in the Community. Master’s Thesis, Graduate Institute of Community Health Care, National Yang Ming Chiao Tung University.

38. Hsu, Yi-hsiung (2020). The Urgent Need to Construct Physical Literacy for Taiwanese Citizens Aged 0 to 100. Sports & Exercise Research, 22(4), i–ii.

39. Hu WY, Chang M, Ke LS. Using SWOT to analyze the promotion of advance care planning-an example of geriatric wards from a medical center. Leadership Nursing. 2017;18(2):13–28.

40. Hu, Chia-Wen. Exploring Learning Styles and Task Adaptation: A Case Study of STEAM Education in Teaching Robotics through Microfilm in Technological Universities. Journal of Teacher Education and Professional Development, 2022, vol. 15, no. 3, pp. 81–117.

41. Huang CC. How Is “Life Education” Possible? Taiwan Journal of General Education. 2019;(24):9–27.

42. Huang, Ching-Mei (2022). The Impact of Self-Regulated Learning Strategy Intervention on the Autonomous Learning Outcomes of Comprehensive High School Students. Master’s Thesis, Department of Industrial Education, College of Technology and Engineering, National Taiwan Normal University.

43. Huang, Jun-Jie (2019). How is “Life Education” Possible? Taiwain Journal of General Education, 24, 9–27.

44. Huang, Kuan-Kai, & Cheng, Po-Wen. (2018). Exploring Doctor-Patient Communication in Hospitals. In National Taipei University of Nursing and Health Sciences, Department of Leisure Industry and Health Promotion (Ed.), Proceedings of the Academic Conference on Leisure Industry and Health Promotion (pp. 119-126). National Taipei University of Nursing and Health Sciences, Department of Leisure Industry and Health Promotion.

45. Huang, Meng-Ming (2020). A Study on the Application of Interactive Media Websites in Teaching Visual Arts Appreciation in Junior High Schools. Master’s Thesis, Art Theory Group, Department of Fine Arts, College of Arts, National Taiwan Normal University.

46. Hung, Pei-Lun & Fang, Chia-Chi. The Role and Impact of Gender Role Stereotypes on Competence and Willingness to Work in Childcare. Taiwan Journal of Counseling Psychology, 2025, vol. 13, no. 1, pp. 65–92.

47. Hung, Yong-Chou. A Brief Discussion on the Trends of Applying Digital Content Materials and Teaching Software in Schools to Enhance Teacher Instruction and Student Learning. Taiwan Education Review Monthly, 2024, 13(5), pp. 70–78.

48. Hung, Yuan-Ming. The Impact of Social Support and Physical, Mental, and Spiritual Health on Life Satisfaction Among the Elderly. Master’s Thesis, Master’s Program in Environment and Development, Dharma Drum Institute of Liberal Arts, 2023.

49. Jan, Fu-Hsuan. An Action Research on the Impact of Experiential Education on Self-Efficacy and Interpersonal Interaction Skills of College Students with Physical and Mental Disabilities. Journal of Special Education in Eastern Taiwan, 2024, No. 27, pp. 147–165.

50. Jiang, Wen-Tzu. Gender Differences in Emotional Expression: A Cross-Situational Analysis. Journal of Educational Psychology, 2018, Vol. 49, No. 3, pp. 345–366.

51. Kuo, Fu-Tzu, Huang, Kuei-Shu, Wang, Cheng-Chung, and Lei, Hsiao-Chuan. An Analysis of Gender Roles in Sports Culture. 2012, Tamkang Sports, 15, pp. 52–56.

52. Lage DE, Caudry DJ, Ackerly DC, Keating NL, Grabowski DC. The care continuum for hospital-ized medicare beneficiaries near death. Annals of Internal Medicine. 2018; 168(10): 748–750.

53. Lai, Chih-Chan. A Review of Current Core Competencies in Life Education: An Examination of “Philosophical Thinking.” Journal of Educational Research, Vol. 70, No. 2, June 2024, pp. 117–154.

54. Lau, Chia-Chi (2019). A Comparative Study of Social Support, Self-Regulated Learning, and Successful Aging Among Older Learners with Different Educational Backgrounds (Unpublished doctoral dissertation). National Kaohsiung Normal University, Kaohsiung City.

55. Lau, Ka-chi (2019). A Comparative Study of Social Support, Self-Regulated Learning, and Successful Aging among Elderly Learners in Different Educational Modalities (Unpublished doctoral dissertation). National Kaohsiung Normal University, Kaohsiung City.

56. Lee FN, Ko MC, Huang SJ, Kao MJ. Palliative and Hospice Care in Emergency Department. Taipei City Medical Journal. 2018;15:61–68.

57. Li, Cheng-Wei, Li, Po-Ang, and Tsai, Ju-Tse. Exploring the Design of Integrating Outdoor Exploration Education into Personal and Social Responsibility Models. Journal of Sports Education in Taiwan, 19(1), 1–28, 2024.

58. Li, Mei-Fang. A Study on the Attitudes of the Elderly Towards Advance Medical Directives: A Case Study of Seniors in a Community Care Center in Yunlin. Master’s Thesis, Department of Thanatology, College of Humanities, Nanhua University, 2021.

59. Li, Pei-fang (2020). Examining the Development of Taiwan’s Community Integrated Care System from the Perspec-tive of Integrated Care. Journal of Community Work and Community Studies, 10(2), 91–134.

60. Li, Pei-Yi. Clarification, Reflection, and Application of Life and Death Education. Counseling and Guidance, No. 227, pp. 2–9, 2004.

61. Liao, Ying-Chin & Kao, Chi-Yin. The Decision-Making Autonomy of Critical Patients at End of Life From the Perspective of Relational Autonomy: A Case Discussion. Nursing Journal, October 2022, Vol. 69, No. 5, pp. 111–119.

62. Lin CP, Peng JK. Patient Autonomy and Advance Care Planning: The United Kingdom Experiences. The Journal of Long-Term Care. 2019; 23(3):169–176.

63. Lin CT, Hsu LY, Wei MR. A Study on the Application of Self-Regulated Learning Cycle Model with Effectiveness of Distance Learning –Taking Adult Lifelong Learning Environment as an Example. Journal of Information Management. 2023;(28):133–154.

64. Lin LH, Chang YC. The Study of Implementation Effectiveness of Senior Learning Centers in Taiwan from the Perspectives of Participants. Journal of Gerontechnology and Service Management. 2019;7(4):363–378.

65. Lin, Chia-Ching, Chen, Chih-Hsien, and Hsieh, Yao-Jen. A Study on the Relationship Between Mental Health Issues, Psychological Needs Satisfaction, and Life Satisfaction Among the Elderly. Taiwan Journal of Counseling Psychology, 2018, Vol. 6, No. 1, 79–109.

66. Lin, Hsin-Yi, Tang, Shu-Chen, Hsieh, Ming-Chuan, He, Hsiu-Ling, Hsu, Ting-Yu, and Chen, Chu-Chieh. A Population-Based Study on Factors Related to Home Death for Patients under Palliative Home Care. Journal of Health Administration, 26(2), 176–201, 2025.

67. Lin, Hsiu-Yi, and Chen, Yu-Ching. A Case Study on Integrating Intergenerational Programs into Interdisciplinary Curricula: A Case of the “Teaching Project on Collective Memory of Islands.” Journal of Welfare Technology and Service Management, Vol. 13, No. 1 (2025).

68. Lin, Hui-Fang. Healthy Practices and Transcendence: Heavy Resistance Training for Frail Elderly in Chiayi’s “Renewal of Withered Trees” Classroom. Master’s Thesis, Department of Applied Sociology, Nanhua University, 2022.

69. Lin, Meng-Chien. (2024). Theory and Practice of Good Death. Master’s and Doctoral Thesis, Graduate Institute of Philosophy, National Central University.

70. Lin, Nai-Hui, and Sun, Kuo-Hua. The Implications, Challenges, and Strategies of Promoting Emotional Education in Universities. Taiwan Educational Review Monthly, October 2021, 5, 101–105.

71. Lin, Shang-Jie, and Wang, Chia-Ling. An Action Research on High School “Ultimate Care” Life Education—Taking aMEI’s “Afterlife” MV as an Example. Journal of Life Education, Vol. 14, No. 2, 2021.

72. Lin, Tzu-Min. Evidence of Life Transformation? Controversies and Reflections on Large-Scale Spiritual Healing Groups. Zhengdao Institute Newsletter, 2025.

73. Lin, Yi-Chun. A Study on the Characteristics, Support Systems, and Learning Methods of Adult Learning. Taiwan Educational Review Monthly, 2023, 12(6), pp. 78–83.

74. Lin, Yu-Chun. Is Lifelong Learning Just Education and Training? Reflecting on the Development of Public Human Resources from the Characteristics of Public Servants. Journal of Public Administration, No. 61, September 2021, pp. 79–114.

75. Liu YC, Liu SM. The Ultimate Concern toward Life Education: Life and Death. The Journal of National Defense University General Education. 2019;(9):149–168.

76. Liu YC. Life Education in Spiritual Meditation. The Journal of National Defense University General Education. 2018;(8):21–36.

77. Liu YF, Ho HH. Reform of Distance Learning Academic Ability Assessments and Follow-Up Guidance Strategies in Primary and Secondary Schools Under the Postpandemic Era. Journal of Education Research. 2023;(346):16.

78. Liu, Yi-Chai & Liu, Shih-Miao (2019). Ultimate Concern of Life Education: Thanatology. The Journal of NDU Gen-eral Education,, 9, 149–168.

79. Lou, C.C., Wang, C.Y., A Study of Constructing a Scale of the Elder Learners’ Social Support, Ming Hsin Journal, 2020/12, 44 (1) Pp. 139–153

80. Ma, Ruei-Chiu, Lin, Pei-Hsuan, Hsiao, Chia-Ying, & Su, Min-Yi (2020). Concept Analysis of Death Competence among Healthcare Providers. Journal of MacKay Nursing, 14(1), 7–13.

81. Mak JM. Clinton M. Promoting a good death: An agenda for outcome research - a review of the literature. Nursing Ethics, 1999;6(2):97–106.

82. Ming-Hao Liang, Hui-Chuan Wei, A Survey on Depression Tendency of Elderly Learners in the Senior Citizens Learning Center, Journal of Health Management; 18(2):45–62

83. Miyashita M, Morita T, Sato K, Hirai K, Shima Y, Uchitomi Y. Good death inventory: A measure for evaluating good death from the bereaved family member’s perspective. Journal of Pain and Symptom Management. 2008; 35(5): 486–498.

84. Miyashita M, Morita T, Sato K, Tsuneto S, Shima Y. A Nationwide Survey of Quality of End-of-Life Cancer Care in Designated Cancer Centers, Inpatient Palliative Care Units, and Home Hospices in Japan: The J-HOPE Study. J Pain Symptom Management. 2015; 50(1): 38–47.

85. O, Cheng-Chun, Wang, Chong-Ying, Zhang, Yu-Tzu, & Hsiao, Chu-Tsen. The Distribution of the Term “Ban Zhuo” in Taiwanese Community News Texts and Recommendations for CSL Cultural Teaching. Chinese Language Teaching Research, 19.3: 43–76, 2022.

86. Pan Su Yu, Lin DYM. The Effect of Auditory Interface on Menu-based Information Navigation for Older Adults. Journal of Ergonomic Study. 2017;19(1):13–27.

87. Steinhauser KE, Christakis NA, Clipp EC, McNeilly M, McIntyre L, Tulsky JA. Factors considered important at the end of life by patients, 123 family, physicians, and other care providers. Journal of American Medical Association. 2000: 284(19): 2476–22482.

88. Su MI, Hsiao CY, Lin PX, Ma JC. Death Literacy Among Health Care Providers: A Concept Analysis. Journal of MacKay Nursing. 2020;14(1):7–13.

89. Sun, Zhi-Chen. How to Learn About Life and Death? Discussing Life and Death Education and Teacher Training for Elderly Life and Death Education. Innovative Care Magazine, October 2024.

90. Sun, Zhi-Chen. The Helplessness of Helpers: A Study of Social Workers Facing Life-and-Death Issues. Master’s Thesis, Department of Thanatology, Nanhua University, 2023.

91. Tang ST. Meanings of dying at home for Chinese patients in Taiwan with terminal cancer. Cancer Nursing. 2000: 23(5); 367–370.

92. Tang ST. When death is imminent: where terminally ill patients with cancer prefer to die and why. Cancer Nursing. 2003: 26(3); 245–251.

93. The Long-Term Care Division of the Ministry of Health and Welfare in Taiwan is actively developing aging policies and smart healthcare, aiming for a significant leap towards all-age inclusiveness and intergenerational well-being, starting from October 2023.

94. Tsai, Chang-Ying. Ethical Dilemmas in End-of-Life Care for Terminal Patients and Ethical Reflection in Social Work. Journal of Contemporary Social Work, 2020, 11(No. 11), pp. 1–29.

95. Tsai, Miao-Chuan (2013). The Impact of Death Education on the Attitudes Toward Death and End-of-Life Planning Among Economically Disadvantaged Elderly Living Alone (Master’s thesis, National Taiwan University).

96. Tseng, Pei-Lu. Discussing the Cross-Cultural Adaptation of School Counseling Teachers. Taiwan Educational Review Monthly, 2021, 10(8), pp. 143–148.

97. Wang CC, Lou CH. Natural Death, Assisted Dying and Criminal Liability. Taiwan Journal of Hospice Palliative Care. 2010;15(1):63–80.

98. Wang, Li-Jing. An Analysis of the Evolution and Development of the Curriculum on Gender Equality Education Issues. Journal of Educational Research (1680-6360), 2025, No. 371, p. 62. DOI: 10.53106/168063602025030371004.

99. Wei, Tsai-Mi. A Study on the Individualization Process of Travel Activities Among Older Learners. Doctoral Dissertation, Department of Social Education, National Taiwan Normal University, 2022.

100. Wu CY, Chen DR, Hung ST. Knowledge and attitudes regarding the Patient Autonomy Act and behavioral intention regarding signing advance decision among in-patients’ family members. Taiwan Journal of Public Health. 2020;39(3):342–349.

101. Wu, Chia-Ying, Chen, Duan-Rong, & Huang, Hsin-Tzu (2020). Family members of hospitalized patients’ awareness, attitudes, and behavioral intentions regarding the Patient Autonomy Act and advance medical decisions. Taiwan Journal of Public Health, 39(3), 342–349.

102. Yang, Jing-Li, Huang, Yu-Shan, & Weng, Kang-Rong. Personal Characteristics, Family Relationships, and Social Participation of Middle-Aged Singles. Demographic Journal, No. 66, June 2023, pp. 75–128.

103. Yang, Shih-Yu, & Cheng, Chang-Hua. Key Strategies for Enhancing Teachers’ Autonomous Learning and Teaching. Taiwan Educational Review Monthly, 2025, 14(5), pp. 129–135.

104. Yeh, Jun-Ting. The Professional Development Journey of Senior Lecturers: Exploration, Growth, and Professional Presentation. Journal of Education, December 2022, No. 59, pp. 119–160.

105. Yen, Pei-Han. An Investigation into the Factors Influencing the Signing of Advance Medical Directives by Caregivers in Tainan Area. Master’s Thesis, Department of Thanatology, Nanhua University, 2024.

106. Ying-Chin Liao, Chi-Yin Kao. The Decision-Making Autonomy of Critical Patients at End of Life From the Perspective of Relational Autonomy: A Case Discussion. October 2022. Nursing Journal, Vol. 69, No. 5, pp. 111–119.

107. You, Hui-Zhen. A Comparative Study of the Views on Life in Christianity and Buddhism. New Century Religious Studies, Vol. 10, No. 2, December 2011, pp. 109–138.

108. Zhu, Mei-Zhen, Chen, Ling-Zhang, and Li, Li-Ling. Discussing Life Education Practices through Gender Issues in Textbooks. 2015, Educational Pulse, No. 3, pp. 53–65.

